# Acceptability of contact management and care of simple cases of COVID-19 at home: a mixed-method study in Senegal

**DOI:** 10.1101/2021.03.10.21253266

**Authors:** Mouhamadou Faly Ba, Valéry Ridde, Amadou Ibra Diallo, Jean Augustin Diégane Tine, Babacar Kane, Ibrahima Gaye, Zoumana Traoré, Emmanuel Bonnet, Adama Faye

## Abstract

**Introduction:** In mid-2020, due to the health system challenges from increased COVID-19 cases, the Ministry of Health and Social Action in Senegal opted for contact management and care of simple cases at home. The study’s objective was to determine the acceptability of contact management, home care of simple cases of COVID-19, and its associated factors.

**Method:** This was a sequential mixed-method study. We collected data from June 11, 2020, to July 10, 2020, for the quantitative survey (N=813) and from August 24 to September 16, 2020, for the qualitative survey (N=30). We carried out a sampling strategy using marginal quotas at the national level. We collected data using a structured questionnaire in a telephone interview for the quantitative survey and using an interview guide formulated from the quantitative survey’s initial results for the qualitative data. We assessed acceptability using binomial logistic regression combined with content analysis.

**Results:** The care of simple cases of COVID-19 at home was well accepted (78.5%). This result was justified for some (saturation of the health system) but not for others (risk of contamination). The use of home contact management was less accepted (51.4%), with risk limitation as the main reason given. The acceptability of home-based care for simple cases was positively associated with knowledge of the modes of transmission of the virus (ORaj: 1.55 [95%CI: 1.04,2.28]), regular research into COVID-19 (ORaj: 2.12 [95%CI: 1.45,3.12]), belief in the existence of treatment (ORaj: 1.82 [95%CI: 1.19,2.83]), and confidence in institutional information (ORaj: 2.10 [95%CI: 1.43,3.10]). The acceptability of home-based contact management was positively associated with knowledge of the modes of transmission of the virus (ORaj: 1.77 [95%CI: 1.27,2.48]), regular research for information on COVID-19 (ORaj: 2.39 [95%CI: 1.76,3.26]), and confidence in the government in the fight against the epidemic (ORaj: 1.51 [95%CI: 1.10,2.08]).

**Conclusion:** Regular information on the disease, knowledge of its mode of transmission and trust in institutions are factors in accepting COVID-19 management at the community level. Authorities should take these factors into account for better communication to improve the acceptability of home-based care.

## INTRODUCTION

To break the chains of transmission of Severe Acute Respiratory Syndrome Coronavirus 2 (SARS-CoV-2), the causative agent of COVID-19, case detection, case management in dedicated centres [1], and screening and quarantine of contacts have been proposed as non-pharmaceutical interventions [2]. These measures aim to prevent the further transmission of secondary infections. They have been used successfully to prevent further outbreaks in South Korea [2].

In Senegal, as soon as the first case of COVID-19 appeared on March 2, 2020, the authorities put a national multi-sectoral action plan in place for monitoring and response [3]. The government accompanied this plan with measures such as border closures, curfews, bans on movement between regions, closure of places of worship, and closure of markets [45]. They established epidemiological treatment centres (ETCs) in all regions to manage COVID cases [19]. On March 22, 2020, they began to isolate contacts in hotel facilities [4].

Despite the unprecedented national measures taken, COVID-19 cases continued to increase (Figure 1) [6,7] leading to an increase in the number of contacts requiring follow-up.

**Figure 1:**
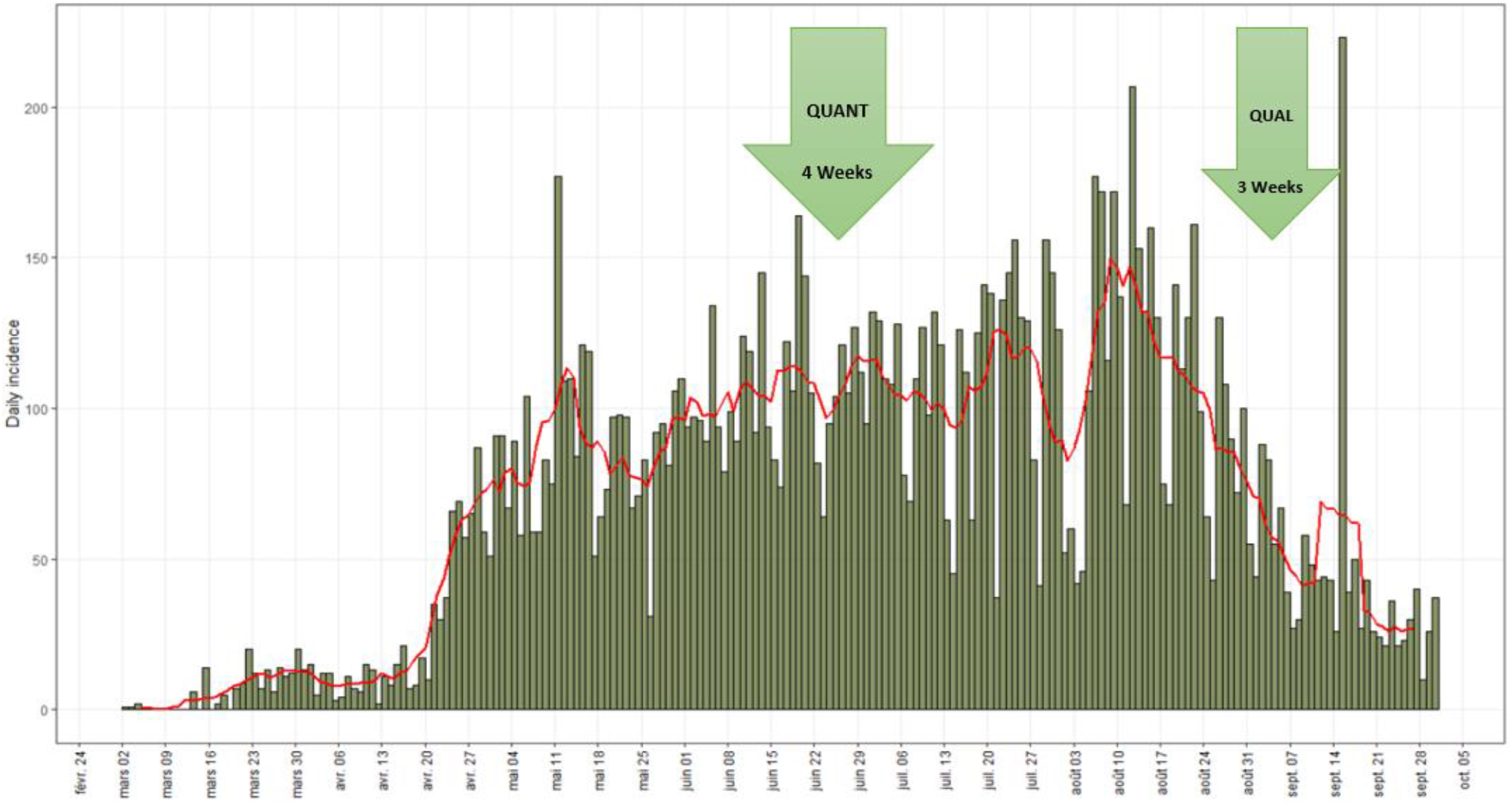
Epidemic curve of COVID-19 confirmed cases (https://covid19datahub.io/)

Faced with this increase in the number numbers and the over-saturation of the health and hotel systems, on May 15, 2020, the authorities decided to stop monitoring contacts in hotels and care of simple cases in the ETCs. For the care of simple cases of COVID-19, they first adopted an extra-hospital care (dedicated sites outside the ETCs) at the end of April 2020 [8] then a home care in July 2020 with protocol to be respected [9]. A simple case is defined as a patient confirmed at COVID-19 who presents signs of uncomplicated upper respiratory tract infection such as fatigue, cough (with or without phlegm), nausea or vomiting, muscle pain, sore throat, nasal congestion, headache, ageusia (loss of taste) and anosmia (loss of smell) [9]. A contact is defined as a person (including caregivers and health workers) who has been exposed to individuals with suspected COVID-19 disease; they are advised to monitor their health for 14 days from the last day of contact [10].

The recent SARS and Ebola epidemics used home-based management approaches [11]. However, there are some risks associated with this strategy, including the spread of the virus within households and in the community [1] and social stigma [12], which undermines these measures’ potential effectiveness. Several countries and the World Health Organization (WHO) have developed guidelines for the home management of COVID cases [10,13-15]. Indeed, the vast majority of patients infected with COVID-19 develop a benign disease [16]. A few studies have shown that the ideal way to control the COVID-19 pandemic is to isolate patients in health facilities with appropriate respiratory precautions, contact tracing, and barrier measures [17-19]. However, isolation in health facilities would result in a shortage of beds for other patients [15]. In this context, home-based management (a familiar environment with family support) is necessary and could help to overcome psychological problems [20]. However, to our knowledge, these guidelines and studies on the subject have not addressed the social acceptability of these measures. Thus, our objective was to determine the acceptability of contact and simple case management of COVID-19 at home and its associated factors in Senegal.

## METHOD

### Study framework

The study took place in the 14 regions of Senegal. The average age in Senegal is 19 years, and males make up 49.7% of the population [21]. The number of confirmed COVID-19 cases on the day the survey started (June 11, 2020) was 4759 [7] with 76.5% of cases in the Dakar region. The organisation of the socio-sanitary sector is pyramidal (central, intermediate and peripheral levels), based on administrative divisions [22]. There are twenty ETCs in all regions of Senegal [23]. They have a capacity of 800 inpatient beds [24] and 80 beds for resuscitation and respirators were available in May throughout the country [25].

### Research estimates, period and study population

It was a mixed-method specification with a sequential explanatory design with a quantitative phase followed by a qualitative phase [26].

We collected quantitative data from June 11 to July 10, 2020, and qualitative data from August 24 to September 16, 2020. The study population consisted of people aged 18 and over in the general population with a mobile phone number.

### Sampling

The quantitative study used a marginal quota sampling strategy [27]. To have a representative sample of the population, we carried out stratification by population weight by region, gender and age group. We randomly generated a nine-digit telephone number list from mobile telephone numbers attributable to Senegal using the Random Digit Dialing (RDD) method. Each number had the same probability of being drawn at random. We integrated this list into a Reactive Auto Dialer (RAD) to trigger calls automatically and optimally. The respondents from the qualitative study were nested in the quantitative study. Their selection followed the same stratification as that of the quantitative sample in order to have a diversity of points of view. A total of 813 individuals took part in the quantitative survey, 30 of whom were in the qualitative survey.

### Data collection

Five interviewers speaking six languages (French, Diola, Wolof, Sérére, Pulaar, Soninké) collected the quantitative data using a structured and closed questionnaire. The interviewers conducted the survey by telephone. They used tablets equipped with Open Data Kit (ODK) software to administer the questionnaire [28,29].

We conceptualised the collected variables in accordance with Bruchon-Schweitzer’s integrative and multifactorial model [30]. This model has good content validity for this study as it integrates most of the variables identified in the literature review. According to the model, we divided the factors in our study into three groups: situational, dispositional, and transactional.

Situational factors are socio-demographic characteristics such as age, gender, region, education level, marital status and economic well-being score. Dispositional factors are knowledge about the cause of the disease, symptoms, modes of transmission and availability of treatment and other variables such as trust in government, information seeking and trust in different information sources (institutions, national media, social networks, health professionals and other applications). Transactional factors are concerns about the epidemic and psychosocial well-being [31]. The independent variables are defined in Appendix 1.

We measured the acceptability (the dependent variable) using a 5-point Likert scale (Strongly agree = 5 not at all agree = 1). It was transformed into binary variables (Yes = Strongly agree and Agree) to determine acceptability levels and identify associated factors.

The qualitative survey was guided by the results of the quantitative analyses. Using a guide formulated from the preliminary descriptive results, the average individual interview duration was 37 minutes. Of the 30 interviews, 28 were conducted in Wolof.

### Data analysis

The quantitative analyses were carried out using R software version 4.0.2. The quantitative variables are described through the mean with its standard deviation and the qualitative variables through the frequencies. We used the Student’s test to compare mean ages, and the Chi2 test to compare other characteristics with a 5% alpha risk. We used binomial logistic regression in the multivariate analysis. We ran two models to determine the factors associated with the acceptability of management of home contacts (model 1) and those associated with care of simple home cases (model 2). We included all variables with p-values below 0.25 in the original models [32]. We used the step-by-step top-down selection procedure in each model to construct the final model. We individually removed variables that did not improve the model. We used the likelihood ratio test compare nested models [33]. We used this multivariate analysis determine adjusted Odds Ratios (ORaj).

For the qualitative data, we transcribed the interviews in full in French and then conducted a manual content analysis [34].

According to the mixed-method approach, divergences and convergences are taken into account in the presentation of the results, particularly concerning the percentages of acceptability of home care measures [35].

### Ethics

The study was approved by the National Ethics Committee for Health Research in Senegal (SEN/20/23).

## RESULTS

The average age of the respondents was 34.70 ± 14.20 years. Males represented 54.6%. In our study, 64.3% of the respondents regularly sought information about COVID-19. The proportion of respondents who trusted institutional information was 56.7% (Appendix 2).

The proportion of participants who accepted care for simple COVID-19 cases at home was 78.5%. In the qualitative survey, some respondents thought that this high percentage was justified by the capacity of the hospitals: “*We had said that the hospitals are full to bursting point, so those with simple cases of COVID, it is better to leave them in t he homes and treat them from there and those who are serious cases are taken to hospital* “(F, 20 years old, Kaolack). For others, this result was astonishing: “*I’m not for that, it’s not safe. Even if the individual has simple COVID-19, it is better to take him and take care of him, otherwise he can contaminate people*” (F, 33, Fatick). Furthermore, 48.6% of the participants did not accept the management of COVID-19 contacts at home. The respondents explained this by a need for risk reduction: “*If you are a contact and the disease is asymptomatic, if you are allowed to be in contact with other people, it can be a risk. So it is better to isolate the person until the incubation period passes. “* (H, 40 years old, Thiès).

The proportion of acceptability of management for home contacts among participants with a good knowledge of the modes of transmission of the virus was 56.1% while those with poor knowledge were 39.0% (p<0.001). The proportion of acceptability of care of simple cases at home among participants who believed that treatment was available was 85.5%, while that of others was 74.8% (p=0.001) (Appendix 3).

Table 1 shows that the acceptability of management for home-based contacts could be based on trust in the government to fight the epidemic (ORaj: 1.51 [95%CI: 1.10,2.08]), knowledge about the modes of transmission of the virus (ORaj: 1.77 [95%CI: 1.27,2.48]), concern about the epidemic (ORaj: 0.68 [95%CI: 0.50,0.93]), and regularly searching for information on COVID-19 (ORaj: 2.39 [95%CI: 1.76,3.26]).

**Table 1:** Results of the multivariate analysis

|  | Acceptability of management<br>for home contacts (Yes) | Acceptability of care for<br>simple home cases (Yes) |
| --- | --- | --- |
| Features | ORaj [95%CI] | ORaj [95%CI] |
| Age | 1,01 [0,99-1,02] | 0,99 [0,98-1,01] |
| **Confidence in the government to**<br>**fight the epidemic** |  |  |
| No | 1 | 1 |
| Yes | **1,51 [1,10-2,08]\*** | 1,30 [0,87-1,94] |
| Features | ORaj [95% CI] | ORaj [95% CI] |
| **Knowledge about the cause of**<br>**COVID-19** |  |  |
| Wrong | 1 | 1 |
| Good | 0,92 [0,66-1,28] | 0,94 [0,62-1,45] |
| **Knowledge of the modes of**<br>**transmission of the virus** |  |  |
| Wrong | 1 | 1 |
| Good | 1,77 [1,27-2,48]* | 1,55 [1,04-2,28]* |
| **Concern about the epidemic** |  |  |
| No | 1 | 1 |
| Yes | 0,68 [0,50-0,93]* | 1,07 [0,73-1,57] |
| **Regularly search for information on**<br>**COVID-19** |  |  |
| No | 1 | 1 |
| Yes | 2,39 [1,76-3,26]* | 2,12 [1,45-3,12]* |
| **Confidence in information from**<br>**social networks** |  |  |
| No |  | 1 |
| Yes |  | 1,11 [0,69-1,85] |
| **Economic well-being score** |  |  |
| Poor |  | 1 |
| Medium |  | 0,69 [0,39-1,21] |
| Rich |  | 0,46 [0,29-0,72]* |
| **Belief in the existence of treatment** |  |  |
| No/NSP |  | 1 |
| Yes |  | 1,82 [1,19-2,83]* |
| **Confidence in institutional**<br>**information** |  |  |
| No/NSP |  | 1 |
| Yes |  | 2,10 [1,43-3,10]* |

The acceptability of care of simple cases at home could be predicted by knowledge of the modes of transmission of the virus (ORaj: 1.55 [95%CI: 1.04,2.28]), regular research of information on COVID-19 (ORaj: 2.12 [95%CI: 1.45,3.12]), wealth based on the score of economic well-being compared to poverty (ORaj: 0.46 [95%CI: 0.29,0.72]), belief in the existence of treatment (ORaj: 1.82 [95%CI: 1.19,2.83]), and trust in institutional information (ORaj: 2.10 [95%CI: 1.43,3.10]).

## DISCUSSION

The study found that while respondents supported care for simple cases of COVID-19 at home, they were more cautious about management for home contacts. These results are interesting given the adoption of this strategy by MoHSA [36]. Highlighted by the qualitative survey, these results can be justified by the fact that the participants are concerned about health system overload, and accept their care at the community level. However, participants are more divided on the management of contacts. WHO recommends isolation of contacts for 14 days after the last exposure with a confirmed case [2]. During the Ebola epidemic, this isolation period was 21 days [37]. A British survey revealed that only 10.9% of contacts adhere to quarantine, and 18.2% adhere to self-isolation [38]. Some of the factors preventing adherence to the isolation may be related to social and financial charges [39]. The 14 days for COVID-19 are difficult to enforce as they take place in the home, and may expose the community to transmission of the virus if people do not isolate themselves. Thus, illustrated by the qualitative survey, these results can be explained by the unknown status of these contacts who may be asymptomatic and then transmit the disease. The experience of the Ebola epidemic in Senegal had shown a negative perception of risk around contacts because people considered them infected with the virus [37]. In addition to strengthening the monitoring of household contacts, efforts should be made to increase people’s understanding of these measures through public health counselling, explaining the importance of ECP of household contacts to reduce transmission, and strong local and social support networks to raise awareness [40].

The study in Senegal found that individuals who trusted institutional sources were more likely to accept care from simple cases at home. Similarly, trust in government in the fight against the epidemic was positively associated with the acceptability of management for home-based contacts. This finding is similar to the study in Israel and China [41] which showed that trust in institutions represented a ‘ reservoir of favourable attitudes and good will‘ during the COVID epidemic [42].

Good knowledge of the modes of transmission of the virus was positively associated with the acceptability of contact management and care of simple cases at home. Since the beginning of the pandemic, the MoHSA has been explaining to the population the importance of respecting collective and individual prevention measures [8]. These prevention measures have been defined as necessary to curb the spread of the virus [43]. Two studies conducted on HIV have shown that individuals with a good knowledge of the modes of transmission of the virus have a better knowledge of the modes of prevention [44,45]. A study conducted in Senegal in April 2020 showed that barrier gestures seemed to be well followed [46]. Compliance with these measures, together with knowledge of how the virus is transmitted, may explain the good acceptability of management among populations.

The regular search for information on COVID-19 was positively associated with the acceptability of contact management and care of simple cases at home. The information provides knowledge that the recipient did not possess or could not foresee [47]. This definition recognises that information as an element of knowledge reduces ignorance about COVID-19. This knowledge will enable the community to consider the extent of the current context and adhere to public health measures. A systematic review showed that the provision of information is an important factor in influencing public acceptability of the authorities’ measures [48]. This leads to a better understanding of the disease and autonomous decision-making in the light of the evolution of the pandemic. This finding seems consistent because people know better what is good for them and are therefore reluctant to accept an intervention that interferes with their own decisions [48].

Belief in the existence of treatment is positively associated with the acceptability of care in simple cases at home. To date, no specific medication is recommended to prevent or treat infection of the new coronavirus [49]. At the beginning of the epidemic, Senegal adopted a treatment protocol based on the treatment of patients with hydroxychloroquine [50] and later with a combination of hydroxychloroquine and azithromycin [51]. This perception of the population in Senegal can be explained by the national communication on the “encouraging” results of this protocol [51], which has become a source of hope in the event of contamination despite the risks involved in its acceptability.

Concern about the epidemic is negatively associated with the acceptability of management for home contacts. Health risks or threats, such as crises, involve emotional connotations and uncertainty about their health and economic implications [52]. These risks would lead to concern about the pandemic which may be due to the prospect of undesirable future consequences [53] and may explain this attitude.

## Limitations

Our study has certain limitations. It only involved people who had a mobile phone, thus excluding marginalised populations. In addition, the cross-sectional nature of the data limits our ability to draw conclusions about causality. However, the sample is representative of the Senegalese population and the use of mixed methods allowed for a better understanding of the results.

## CONCLUSION

This study shows that being regularly informed about the disease, knowing how it is transmitted, and trusting institutions are important factors in the acceptance of COVID-19 management at the community level. It will be important for the authorities to consider and integrate these aspects for a more effective strategy. However, it is also necessary to have messages that are adapted and targeted according to the categories of the population.

## Data Availability

Data are available upon request to the authors.

## ACKNOWLEDGEMENTS

We would like to thank the five interviewers who participated in the data collection: Tabaski Diouf, Coumba Sow, Fatoumata Dieme, Rokhaya Gueye and Mafoudya Camara. To our technical partner Cloudlyyours, who have been able to put in place all the necessary tools for data collection.

## CONFLICT OF INTEREST

The authors have no conflicts of interests regarding the publication of this paper.

## DATA AVAILABILITY STATEMENT

Data are available upon request to the authors.

## FUNDING

This research was conducted as part of the ARIACOV programme (Appui à la Riposte Africaine à l’épidémie de Covid-19), which receives funding from the French Development Agency through the “COVID-19 - Santé en commun” initiative.

## Appendices

### Appendix 1: Definition of independent variables

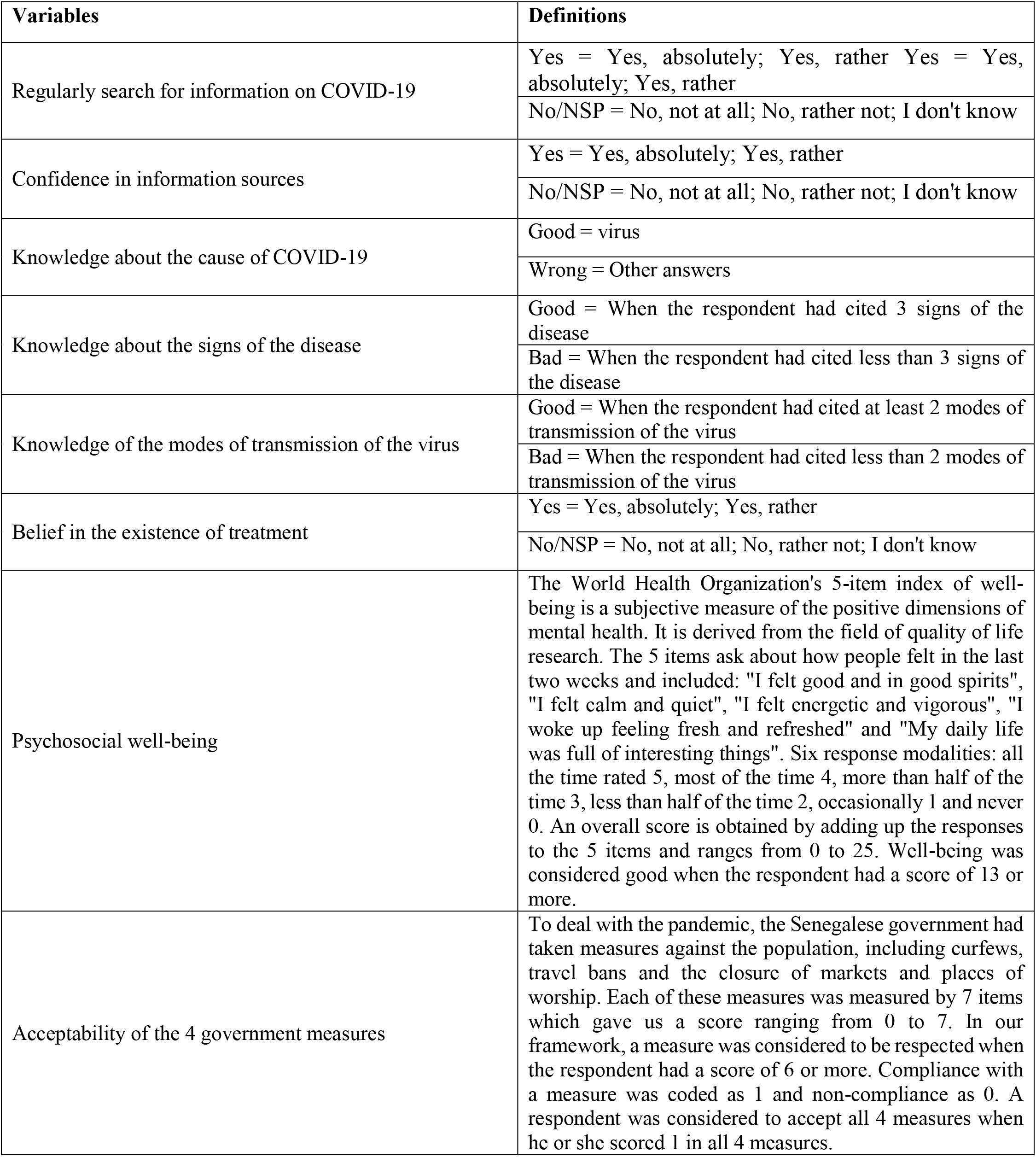

### Appendix 2: Distribution of respondents by characteristics (N=813)

| VARIABLES | n (%) |
| --- | --- |
| **Sex** |  |
| Woman | 369 (45.4%) |
| Man | 444 (54.6%) |
| **Region** |  |
| Dakar | 247 (30.4%) |
| Outside Dakar | 566 (69.6%) |
| **Level of education** |  |
| Uneducated | 346 (42.6%) |
| Primary | 154 (18.9%) |
| Secondary/Higher | 313 (38.5%) |
| **Marital status** |  |
| Married | 499 (61.4%) |
| Unmarried | 314 (38.6%) |
| **Economic well-being score** |  |
| Poor | 229 (28.2%) |
| Medium | 165 (20.3%) |
| Rich | 419 (51.5%) |
| **Confidence in the government to fight the epidemic** |  |
| No | 524 (64.5%) |
| Yes | 289 (35.5%) |
| **Regular search for information on COVID-19** |  |
| No/NSP | 290 (35.7%) |
| Yes | 523 (64.3%) |
| **Confidence in institutional information** |  |
| No/NSP | 352 (43.3%) |
| Yes | 461 (56.7%) |
| **Confidence in national media information** |  |
| No/NSP | 135 (16.6%) |
| Yes | 678 (83.4%) |
| **Confidence in information from social networks** |  |
| No/NSP | 634 (78.0%) |
| Yes | 179 (22.0%) |
| **Confidence in information from health professionals** |  |
| No/NSP | 56 (6.9%) |
| Yes | 757 (93.1%) |
| **Confidence in information from WhatsApp or other application** |  |
| No/NSP | 653 (80.3%) |
| Yes | 160 (19.7%) |
| **Knowledge about the cause of COVID-19** |  |
| Wrong | 593 (72.9%) |
| Good | 220 (27.1%) |
| **Knowledge about the signs of the disease** |  |
| Wrong | 595 (73.2%) |
| Good | 218 (26.8%) |
| **Knowledge of the modes of transmission of the virus** |  |
| Wrong | 223 (27.4%) |
| Good | 590 (72.6%) |
| **Belief in the existence of treatment** |  |
| No/NSP | 531 (65.3%) |
| Yes | 282 (34.7%) |
| **Psychosocial well-being** |  |
| Wrong | 41 (5.0%) |
| Good | 772 (95.0%) |
| **Concern about the epidemic** |  |
| No | 442 (54.4%) |
| Yes | 371 (45.6%) |
| **Acceptability of management for home contacts** |  |
| No/NSP | 395 (48.6%) |
| Yes | 418 (51.4%) |
| **Acceptability of care for simple cases at home** |  |
| No/NSP | 175 (21.5%) |
| Yes | 638 (78.5%) |

### Appendix 3: Breakdown of respondents by characteristics and acceptability of home -based care (N=813) put the tables in appendices

| VARIABLES | N (% Acceptability of management of home contacts) | p-value | N (% Acceptability of care for simple cases at home) | p-value |
| --- | --- | --- | --- | --- |
| **Age ($\mu \pm \sigma$)** | 35.5 (14.2) | 0.097 | 34.6 (14.1) | 0.653 |
| **Sex** |  | 0.754 |  | 0.854 |
| Woman | 369 (50.7%) |  | 369 (78.0%) |  |
| Man | 444 (52.0%) |  | 444 (78.8%) |  |
| **Level of education** |  | 0.594 |  | 0.056 |
| Uneducated | 346 (50.0%) |  | 346 (74.6%) |  |
| Primary | 154 (50.0%) |  | 154 (79.9%) |  |
| Secondary/Higher | 313 (53.7%) |  | 313 (82.1%) |  |
| **Region** |  | 0.492 |  | 0.155 |
| Outside Dakar | 566 (50.5%) |  | 566 (77.0%) |  |
| Dakar | 247 (53.4%) |  | 247 (81.8%) |  |
| **Marital status** |  | 1.000 |  | 0.588 |
| Unmarried | 314 (51.3%) |  | 314 (79.6%) |  |
| Married | 499 (51.5%) |  | 499 (77.8%) |  |
| **Economic well-being score** |  | 0.570 |  | **0.035** |
| Poor | 229 (53.7%) |  | 229 (83.8%) |  |
| Medium | 165 (52.7%) |  | 165 (79.4%) |  |
| Rich | 419 (49.6%) |  | 419 (75.2%) |  |
| **Confidence in the government to fight the epidemic** |  | 0.081 |  | 0.309 |
| No | 524 (49.0%) |  | 524 (77.3%) |  |
| Yes | 289 (55.7%) |  | 289 (80.6%) |  |
| **Regular search for information on COVID-19** |  | **<0.001** |  | **<0.001** |
| No/NSP | 290 (35.2%) |  | 290 (66.2%) |  |
| Yes | 523 (60.4%) |  | 523 (85.3%) |  |
| **Confidence in institutional information** |  | **0.004** |  | **<0.001** |
| No/NSP | 352 (45.5%) |  | 352 (69.6%) |  |
| Yes | 461 (56.0%) |  | 461 (85.2%) |  |
| **Confidence in national media information** |  | 0.355 |  | 0.088 |
| No/NSP | 135 (47.4%) |  | 135 (72.6%) |  |
| Yes | 678 (52.2%) |  | 678 (79.6%) |  |
| **Confidence in information from social networks** |  | **<0.001** |  | 0.148 |
| No/NSP | 634 (47.8%) |  | 634 (77.3%) |  |
| Yes | 179 (64.2%) |  | 179 (82.7%) |  |
| **Confidence in information from health professionals** |  | **0.636** |  | **0.410** |
| No/NSP | 56 (55.4%) |  | 56 (73.2%) |  |
| Yes | 757 (51.1%) |  | 757 (78.9%) |  |
| **Confidence in information from WhatsApp or other application** |  | **0.001** |  | **0.289** |
| No/NSP | 653 (48.5%) |  | 653 (77.6%) |  |
| Yes | 160 (63.1%) |  | 160 (81.9%) |  |
| **Knowledge about the cause of COVID-19** |  | **0.826** |  | **0.869** |
| Wrong | 593 (51.1%) |  | 593 (78.2%) |  |
| Good | 220 (52.3%) |  | 220 (79.1%) |  |
| **Knowledge about the signs of the disease** |  | **0.034** |  | **0.935** |
| Wrong | 595 (49.1%) |  | 595 (78.3%) |  |
| Good | 218 (57.8%) |  | 218 (78.9%) |  |
| **Knowledge of the modes of transmission of the virus** |  | **<0.001** |  | **0.002** |
| Wrong | 223 (39.0%) |  | 223 (70.9%) |  |
| Good | 590 (56.1%) |  | 590 (81.4%) |  |
| **Belief in the existence of treatment** |  | **0.065** |  | **0.001** |
| No/NSP | 531 (49.0%) |  | 531 (74.8%) |  |
| Yes | 282 (56.0%) |  | 282 (85.5%) |  |
| **Concern about the epidemic** |  | **0.022** |  | **0.651** |
| No | 442 (55.2%) |  | 442 (79.2%) |  |
| Yes | 371 (46.9%) |  | 371 (77.6%) |  |
| **Psychosocial well-being** |  | **0.156** |  | **0.605** |
| Wrong | 41 (63.4%) |  | 41 (82.9%) |  |
| Good | 772 (50.8%) |  | 772 (78.2%) |  |

## Notes

### Competing Interest Statement

The authors have declared no competing interest.

### Funding Statement

This research was conducted as part of the ARIACOV programme (Appui a la Riposte Africaine a l'epidemie de Covid-19), which receives funding from the French Development Agency through the "COVID-19 - Sante en commun" initiative.

### Author Declarations

The research was accepted by Senegal's national health research ethics committee (SEN/20/23). Phone: (+221) 33 556 89 56

## REFERENCES

1. Wilder-Smith A, Cook AR, Dickens BL. Institutional versus home isolation to curb the COVID-19 outbreak-Authors’ reply. Lancet [Internet]. 2020 Nov 21 [cited 2020 Dec 17];(396(10263):1632–3. Available from: https://www.thelancet.com/journals/lancet/article/PIIS0140-6736(20)32171-1/fulltext

2. Quilty BJ, Clifford S, Hellewell J, Russell TW, Kucharski AJ, Flasche S, et al. Quarantine and testing strategies in contact tracing for SARS -CoV-2: a modelling study. Lancet Public Heal [Internet]. 2021 Jan [cited 2021 Jan 23];1–9. Available from: https://linkinghub.elsevier.com/retrieve/pii/S246826672030308X

3. Government of Senegal. Council of Ministers of 04 March 2020 [Internet]. 2020 [cited 2021 Jan 24]. Available from: https://www.sec.gouv.sn/actualité/conseil-des-ministres-du-04-mars-2020

4. Emergency Health Operations Centre. Situation Report No. 40 of 23 July 2020 [Internet]. Dakar, Senegal; 2020. Available from: http://www.sante.gouv.sn/sites/default/files/SITREP_40_COVID_SNpdf.

5. Republic of Senegal. CORONAVIRUS : President Macky Sall takes several steps [Internet]. 2020 [cited 2020 Nov 18]. Available from: http://www.mesr.gouv.sn/index.php/2020/03/15/coronavirus-le-president-de-la-republique-macky-sall-prend-plusieurs-mesures/

6. Ministry of Health a nd Social Action. Press release 22 of 23 03 2020 [Internet]. Dakar, MSAS. 2020 [cited 2020 Sep 10]. p. 1. Available from: http://www.sante.gouv.sn/sites/default/files/COMMUNIQUE22of23March2020.pdf

7. Ministry of Health and Social Action. Press release 222 of 09 10 2020 [Internet]. Dakar, MSAS. 2020 [cited 2020 Oct 9]. p. 2. Available from: http://www.sante.gouv.sn/sites/default/files/Communiqué222du09102020.pdf

8. Ministry of Health and Social Action. Pandemic COVID-19 / SENEGAL COMMUNIQUE 67 [Internet]. Dakar, Senegal; 2020. p. 2. Available from: http://www.sante.gouv.sn/sites/default/files/COMMUNIQUE67DU07MAI2020.pdf

9. Emergency Health Operations Centre. Standard Operating Procedure (SOP) : management of simple cases at home in the context of COVID-19. Dakar, Senegal; 2020.

10. World Health Organization. Home-based care for COVID-19 patients with mild symptoms, and management of their contacts [Internet]. Geneva, Switzerland; 2020. Available from: https://apps.who.int/iris/bitstream/handle/10665/331527/WHO-nCov-IPC-HomeCare-2020.3-fre.pdf?sequence=1&isAllowed=y

11. Faye SL. The “exceptionality” of Ebola and popular “reticence” in Guinea -Conakry. Reflections from a symmetrical anthropological approachThe “exceptionality” of Ebola and popular “reticences” in Guinea -Conakry. Reflections from a symmetrical ant. Anthropol Santé [Internet]. 2015 Nov 25 [cited 2021 Feb 7];(11). Available from: http://journals.openedition.org/anthropologiesante/1796

12. Syed Mohammed. 94% of COVID patients treated in isolation at home, according to a survey - The Hindu [Internet]. The Indu. 2020 [cited 2020 Dec 17]. Available from: https://www.thehindu.com/news/cities/Hyderabad/94-covid-patients-treated-in-home-isolation-shows-survey/article32663377.ece

13. Gouvernement Australien. Home isolation guidance when unwell (suspected or confirmed cases) [Internet]. Canberra, Australie; 2020 [cited 2020 Dec 17]. Available from: https://www.health.gov.au/sites/default/files/documents/2020/03/coronavirus-covid-19-information-about-home-isolation-when-unwell-suspected-or-confirmed-cases.pdf

14. Canadian government. Available at jour : Public health management of COVID-19 cases and associated contacts -Canada.ca [Internet]. 2020 [cited 2020 Dec 17]. Available from: https://www.canada.ca/fr/sante-publique/services/maladies/2019-nouveau-coronavirus/professionnels-sante/directives-provisoires-cas-contacts.html

15. Ayaz CM, Dizman GT, Metan G, Alp A, Unal S. Out -patient management of patients with COVID-19 on home isolation. Infez Med. 2020;(28(3):351 –6.

16. INSERM. Coronavirus and Covid-19 [Internet]. 2020 [cited 2021 Jan 8]. Available from: https://www.inserm.fr/information-en-sante/dossiers-information/coronavirussars-cov-et-mers-cov

17. Dickens BL, Koo JR, Wilder-Smith A, Cook AR. Institutional, not home -based, isolation could contain the COVID-19 outbreak. Lancet [Internet]. 2020;(395(10236):1541 –2. Available from: http://dx.doi.org/10.1016/S0140-6736(20)31016-3

18. Kucharski AJ, Klepac P, Conlan AJK, Kissler SM, Tang ML, Fry H, et al. Effectiveness of isolation, testing, contact tracing, and physical distancing on reducing transmission of SARS-CoV-2 in different settings: a mathematical modelling study. Lancet Infect Dis. 2020;(20(10):1151–60.

19. MacIntyre CR. Case isolation, contact tracing, and physical distancing are pillars of COVID-19 pandemic control, not optional choices. Lancet Infect Dis [Internet]. 2020;(20(10):1105–6. Available from: http://dx.doi.org/10.1016/S1473-3099(20)30512-0

20. Organisation Mondiale de la Santé. Home care for patients with suspected or confirmed COVID-19 and management of their contacts [Internet]. World Health Organization. Genève, Suisse; 2020. Available from: https://www.who.int/publications-detail/home-care-for-patients-with-suspected-novel-coronavirus-(ncov)-infection-presenting-with-mild-symptoms-and-management-of-contacts

21. National Agency for Statistics and Demography [Internet]. 2020 [cited 2020 Nov 19]. Available from: http://www.ansd.sn/

22. Ministry of Health and Social Action. Senegal: Plan National de Développement Sanitaire et Social (PNDSS) [Internet]. Dakar, Senegal; 2019. Available from: http://www.sante.gouv.sn/sites/default/files/1MSASPNDSS20192028FinalVersion.pdf

23. Emergency Health Operations Centre. Situation Report No. 78 of 18 January 2021 [Internet]. Dakar, Senegal; 2020. Available from: http://www.sante.gouv.sn/sites/default/files/SITREP78Covid-1918-01-2021.pdf

24. Cissé M. Senegal. Covid-19: The number of beds increased as a matter of urgency in the face of the worrying development of the pandemic [Internet]. Le360 Africa. 2020 [cited 2021 Jan 24]. Available from: https://afrique.le360.ma/senegal/societe/2020/05/01/30412-senegal-covid-19-le-nombre-de-lits-augmente-en-urgence-face-levolution-inquietante-de-la-pandemie

25. Marbot O. Number of resuscitation beds and respirateurs : What is the status of Afrique? [Internet]. Young Africa. 2020 [cited 2021 Jan 24]. Available from: https://www.jeuneafrique.com/924087/societe/nombre-de-lits-de-reanimation-et-de-respirateurs-ou-en-est-lafrique/

26. Greene J. Toward a methodology of mixed methods social inquiry. Res Sch. 2006;(13(1):93–8.

27. Deville J-C. A theory of quota surveys.pdf. Survey Tech, Stat Canada. 1991;(12(2):177 – 95.

28. Pigeon-Gagné É, Hassan G, Yaogo M, Ridde V. An exploratory study assessing psychological distress of indigents in Burkina Faso: a step forward in understanding mental health needs in West Africa. Int J Equity Health. 2017;(16(1):143.

29. Ouédraogo S, Ridde V, Atchessi N, Souares A, Koulidiati JL, Stoeffler Q, et al. Characterisation of the rural indigent population in Burkina Faso: A screening tool for setting priority healthcare services in sub -Saharan Africa. BMJ Open. 2017;(7(10):9–11.

30. Directorate of Research, Evaluation Studies and Statistics (DREES). Acceptability of the main types of serious adverse events associated with care in the general population and among doctors [Internet]. Paris, France; 2011. Available from: https://drees.solidarites-sante.gouv.fr/IMG/pdf/serieetud108.pdf

31. Organisation Mondiale de la Santé. Wellbeing Measures in Primary Health Care/ The Depcare Project [Internet]. Report on a WHO Meeting. Genève, Suisse; 1998. Available from: http://www.euro.who.int/data/assets/pdf_file/0016/130750/E60246.pdf

32. Hosmer DW, Lemeshow S. Applied Logistic Regression. 2nd Editio. Wiley-Interscience Publication. New York, USA; 2000. 392 p.

33. Zhang Z. Model building strategy for logistic regression: Purposeful selection. Ann Transl Med. 2016;(4(6):4–10.

34. Miles MB, Huberman AM. Qualitative Data Analysis. Second Edition. Thousand Oaks : SAGE Publications. 1994. 354 p.

35. Pluye P, Grad RM, Levine A, Nicolau B. Understanding divergence of quantitative and qualitative data (or results) in mixed methods studies. Int J Mult Res Approaches. 2009;(3(1):58–72.

36. Emergency Health Operations Centre. Situation Report No. 35 of 06 July 2020 [Internet]. Dakar, Senegal; 2020. Available from: http://www.sante.gouv.sn/sites/default/files/SITREP_35_COVID_SN.pdf

37. Desclaux A, Badji D, Ndione AG, Sow K. Accepted monitoring or endured quarantine? Ebola contacts’ perceptions in Senegal. Soc Sci Med [Internet]. 2017 178:38–45. Available from: http://dx.doi.org/10.1016/j.socscimed.2017.02.009

38. Smith LE, Potts HW, Amlôt R, Fear NT, Michie S, James Rubin G. Adherence to the test, trace and isolate system: results from a time series of 21 nationally representative surveys in the UK (the COVID-19 Rapid Survey of Adherence to Interventions and Responses [CORSAIR] study). medRxiv [Internet]. 2020 [cited 2021 Feb 7];1–47. Available from: https://doi.org/10.1101/2020.09.15.20191957

39. Webster RK, Brooks SK, Smith LE, Woodland L, Wessely S, Rubin GJ. How to improve adherence with quarantine: rapid review of the evidence. Public Health. 2020 May 1 182:163–9.

40. Webster RK, Brooks SK, Smith LE, Woodland L, Wessely S, Rubin GJ. How to improve adherence with quarantine: rapid review of the evidence. Public Health [Internet]. 2020 May 1 [cited 2021 Jan 23]182:163–9. Available from: /pmc/articles/PMC7194967/?report=abstract

41. Liu XJ, Mesch GS. The adoption of preventive behaviors during the COVID-19 pandemic in China and Israel. Int J Environ Res Public Health. 2020; 17(19):1–18.

42. Guglielmi S, Dotti Sani GM, Molteni F, Biolcati F, Chiesi AM, Ladini R, et al. Public acceptability of containment measures during the COVID-19 pandemic in Italy: how institutional confidence and specific political support matter. Int J Sociol Soc Policy. 2020;

43. World Health Organization. COVID-19 Strategy Update [Internet]. Geneva, Switzerland; 2020 [cited 2021 Jan 24]. Available from: https://www.who.int/docs/default-source/coronaviruse/strategy-update-french.pdf

44. Ibrahim AA, Ali KM, Mohammed AE. Knowledge, counseling and test acceptability regarding AIDS among Knowledge, counseling and test acceptability regarding AIDS among secondary school students, Khartoum, Sudan. 2017;(October).

45. Hesse E. Knowledge about the transmission of the AIDS virus [Internet]. Brussels, Belgium; 2008. Available from: https://his.wiv-isp.be/fr/Documentspartages/HI_EN_2008.pdf

46. Nestour A Le, Samba M, Laura M. Telephone survey on the Covid crisis in Senegal. 2020.

47. Tchouassi G. Information needs in companies. Vol. 24, Revue Congolaise de Gestion. 2017. 63 p.

48. Diepeveen S, Ling T, Suhrcke M, Roland M, Marteau TM. Public acceptability of government intervention to change health-related behaviours: A systematic review an d narrative synthesis. BMC Public Health [Internet]. 2013 Dec 15 [cited 2021 Feb 7];(13(1):756. Available from: http://bmcpublichealth.biomedcentral.com/articles/10.1186/1471-2458-13-756

49. World Health Organization. COVID-19 : What you need to know [Internet]. World Health Organization. 2020 [cited 2020 Nov 18]. Available from: https://www.who.int/fr/emergencies/diseases/novel-coronavirus-2019/question-and-answers-hub/q-a-detail/q-a-coronaviruses?gclid=EAIaIQobChMInP6JweTF7AIVTPlRCh34kgtjEAAYASAAEgJe0vD_BwE

50. Emedia. Pr Seydi: “Les chloroquine results are absolutely encourageants” [Internet]. Emedia. 2020 [cited 2020 Nov 19]. Available from: http://emedia.sn/VIDEO-PR-SEYDI-LES-RESULTATS-DE-LA-CHLOROQUINE-SONT-ABSOLUMENT-ENCOURAGEANTS.html

51. Emedia. Chloroquine & Azythromicine : Prof. Seydi revalidates the “Raoult protocol” [Internet]. Emedia. 2020 [cited 2020 Nov 19]. Available from: http://www.emedia.sn/CHLOROQUINE-AZYTHROMICINE-Pr-SEYDI-VALIDE-A-NOUVEAU-LE-PROTOCOLE-DE-RAOULT.html

52. Faye M, Diatta JS. The Senegalese Government’s communication to the COVID-19 test. Akofena [Internet]. 2020 Oct [cited 2021 Jan 9];255–66. Available from: http://revue-akofena.org/wp-content/uploads/2020/10/20-T03-34-pp.-255-266.pdf

53. Quenum GGY, Ertz M. Adaptation strategies in consumption during the COVID crisis - 19 : what hope for consumption responsable? Organ Territ. 2020;(29(3):87–9.

